# Working Memory Gating in Obesity: Insights from a Case-Control fMRI Study

**DOI:** 10.1101/2023.09.13.23295435

**Authors:** Nadine Herzog, Hendrik Hartmann, Lieneke K. Janssen, Maria Waltmann, Lorenz Deserno, Sean J. Fallon, Annette Horstmann

## Abstract

Computational models and neurophysiological data propose that a ‘gating mechanism’ coordinates distractor-resistant maintenance and flexible updating of working memory contents: While maintenance of information is mainly implemented in the prefrontal cortex, updating of information is signaled by phasic increases in dopamine in the striatum. Previous literature demonstrates structural and functional alterations in these brain areas, as well as differential dopamine transmission among individuals with obesity, suggesting potential impairments in these processes. To test this hypothesis, we conducted an observational case-control fMRI study, dividing participants into groups with and without obesity based on their BMI. We probed maintenance and updating of working memory contents using a modified delayed match to sample task and investigated the effects of SNPs related to the dopaminergic system. While the task elicited the anticipated brain responses, our findings revealed no evidence for group differences in these two processes, neither at the neural level nor behaviorally. However, depending on Taq1A genotype, which affects dopamine receptor density in the striatum, participants with obesity performed worse on the task. In conclusion, this study does not support the existence of overall obesity-related differences in working memory gating. Instead, we propose that potentially subtle alterations may manifest specifically in individuals with a ’vulnerable’ genotype.

## 1. INTRODUCTION

In order to act in a goal-directed manner, we have to flexibly adapt to an ever-changing environment. While, in some situations, we need to maintain information relevant to our goals and ignore distractors, in other situations we might need to flexibly update information whenever new goal-relevant information becomes available. A cognitive system enabling such adaptation is working memory (WM). Computational models and neurophysiological data propose that the coordination between distractor-resistant maintenance and flexible updating of WM contents is achieved via a so-called ‘gating mechanism’: While distractor-resistant maintenance of WM representations is mainly implemented in the prefrontal cortex, updating of those representations is achieved via phasic increases in dopamine in the striatum (Durstewitz and Seamans, 2008, O’Reilly and Frank, 2006, Badre & Frank 2012, Chatham & Badre 2013, Frank & Badre 2012). Notably, decreases in tonic dopamine in the striatum have the opposite effect, setting the threshold for updating signals higher, thus impeding updating but probably easing ignoring (O’Reilly and Frank, 2006). One’s ability to ignore or update (ir)relevant information, should hence vary according to individual dopamine levels (e.g. Cools & D’Esposito, 2011; Furman et al., 2021, Jongkees, 2020). Crucially, there is evidence that preeminence in one process may come at the expense of the other (Fallon and Cools, 2017).

In individuals with obesity, the accurate adaptation of WM representations seems to be compromised. There are several studies pointing toward generally diminished WM functioning in obesity (Yang, et al., 2020, 2019, 2018; Gonzales et al., 2010; Coppin et al., 2014), but also others pointing toward more specific impairments in cognitive adaptation, which requires updating of WM contents (e.g. Huang et al., 2019). For example, participants with obesity have been shown to prefer short-term rewards despite negative long-term outcomes (Horstmann et al., 2011; Mathar et al., 2015) or to repeat previously rewarded actions, despite current devaluation (Horstmann et al., 2015a). These findings suggest that obese individuals might fail to update following negative consequences of their actions, which could contribute to dysfunctional preservation of maladaptive (eating-) behaviors. In line with this, it has been suggested that (moderate) obese individuals (30 kg/m² < BMI < 45 kg/m²) have a decreased dopamine tone in the striatum (Horstmann et al., 2015b). Additionally, functional and structural changes in the above-mentioned WM-related brain areas have been observed in obesity (Wang et al., 2001; Dunn et al., 2012; Eisenstein et al., 2013; Kessler et al., 2014; Cosgrove et al., 2015; Volkow et al., 2008, 2011; Horstmann et al., 2011; Mathar et al., 2017).

Together these findings suggest that working memory updating might be compromised in individuals with obesity. Until today, however, it has not been tested systematically which of the two sub-processes contribute to the observed WM impairments. In the present study, we, therefore, aim to fill this gap and elucidate the role of the specific WM sub-processes in obesity.

To this end, we implemented an observational case-control fMRI-based study. We grouped participants based on their BMI (lean: 18,5 - 25 kg/m²; obese: 30-45 kg/m²) and let them do a modified version of a delayed match-to-sample task (Fallon and Cools, 2014; Fallon et al, 2017) in an MRI Scanner. This task comprises specific conditions probing updating and distractor-resistant maintenance of mental representations. Given the observed behavioral and putative dopaminergic differences in obesity, we expect obese participants to display worse WM updating (probably easing more rigid distractor resistance) when compared to lean controls. Furthermore, we expect these differences to be represented in condition-specific changes in brain activity, where activity in the PFC will be associated with ignoring distractors and activity in the striatum with updating of representations.

On a more exploratory basis, we aim to investigate the impact of two single-nucleotide polymorphisms (SNPs), COMT and Taq1A, which have been related to frontal and striatal dopamine, respectively (Käenmäki et al., 2010; Syvänen et al., 1997; Hirvonen et al., 2005) and to working memory gating (Furman et al., 2020). We speculate that the potential differences in maintenance and updating between obese and lean participants might be influenced by COMT Val158Met and Taq1A genotype.

## 2. MATERIALS AND METHODS

### 2.1. Participants

As part of a larger study protocol (see 10.17605/OSF.IO/FYN6Q), 78 participants were recruited via the internal participant database of the Max Planck Institute for Human Cognitive and Brain Sciences (Leipzig, Germany) and via advertisements in public places and university facilities. Participants were specifically screened to exclude a history of clinical drug or alcohol abuse, neurological or psychiatric disorders, or first-degree relative history of neurological or psychiatric disorders. None of the participants had symptoms of depression, as assessed via a screening interview using the Structured Clinical Interview for DSM-IV (SCID, Wittchen 1997). This study was conducted in compliance with the principles of the Declaration of Helsinki and was authorized by the Ethics Committee of the Medical Faculty at the University of Leipzig (385/17-ek). All participants provided written informed consent before participation and were compensated for their time.

### 2.2. Study design

Participants were asked to come to the lab on two occasions. Firstly, for a screening session where in- and exclusion criteria were checked. We measured weight and height to calculate BMI and assign participants to the respective group (obese = 30 kg/m² < BMI < 45 kg/m²; lean = 18,5 kg/m² < BMI < 25 kg/m²). After inclusion, participants first underwent a blood draw to assess COMT Val158Met and Taq1A genotype. After the blood draw, participants completed a battery of neuropsychological tests including the Trail Making Test (TMT) A and B (Reitan, R. M., 1955) to test executive functioning, the Digit Symbol Substitution Test (DSST) to assess processing speed (Wechsler, 1997), Digit Span to test baseline working memory functioning, (Wechsler, 1997), and the German “Wortschatztest” (Lehrl, 2005) as a proxy for (verbal) IQ. Subsequently, two more behavioral tasks were performed (described somewhere else), and participants filled in several questionnaires assessing eating behavior, clinical symptoms, personality traits, and impulsiveness (see Table S1 in supplementary materials). On the second test day, participants first completed two tasks during a 3T MRI scan, one of which was the WM task investigated in this study. The two tasks were counterbalanced in order. After the MRI scan, participants completed a third behavioral task. The exact study design can be found in the larger study protocol mentioned above. The study-specific pre-registration can be found here: 10.17605/OSF.IO/6WRM2.

### 2.3 Working Memory Task

Participants performed a modified version of a delayed match-to-sample task originally designed by Fallon and Cools (2014). In this task, participants had to remember two target stimuli, signaled by the letter “T” centered between the two stimuli, and later evaluate whether one of the target stimuli matches a presented probe or not. The task consists of four task conditions: update, ignore, and two retention-interval matched control conditions (control short and control long). In the update condition, participants are first presented with two target stimuli (signaled by a centered “T”). Next, they see a new pair of target stimuli (again indicated by a centered “T”) that replace the previously presented stimuli as target. During the subsequent probe phase, participants have to match the probe with the two most recently presented target stimuli by pressing either the left or the right button, thereby choosing “yes” (it is a match) or “no” (it is not a match) respectively. In the ignore condition, participants are first presented with a set of target stimuli (centered “T”) again. After the first set of target stimuli, however, they are presented with two new stimuli that are marked as non-targets (centered “N”). This time, the two new non-target stimuli have to be ignored and the previous target stimuli have to be maintained and matched to the probe. The two control conditions do not have any interference and are matched to the temporal delay between encoding the to-be-matched targets and the probe in the update and ignore condition. The probe is presented for 2000 msec in which participants have to respond. The task is separated into four blocks, with feedback on overall performance presented after each of those blocks, providing a short break for the participant. The four task conditions were counterbalanced among all blocks. Each block consists of 32 trials, amounting to 128 trials in total. Each trial is separated by a jittered inter-trial interval ranging from 2000 to 6000 msec. The stimuli are randomly computer-generated, monochromatic RGB “spirographs”. Outcome measures are accuracy and reaction time. The total duration of the task is approximately 30 minutes. A systematic outline of the task is depicted in Figure 1.

**Figure 1.**
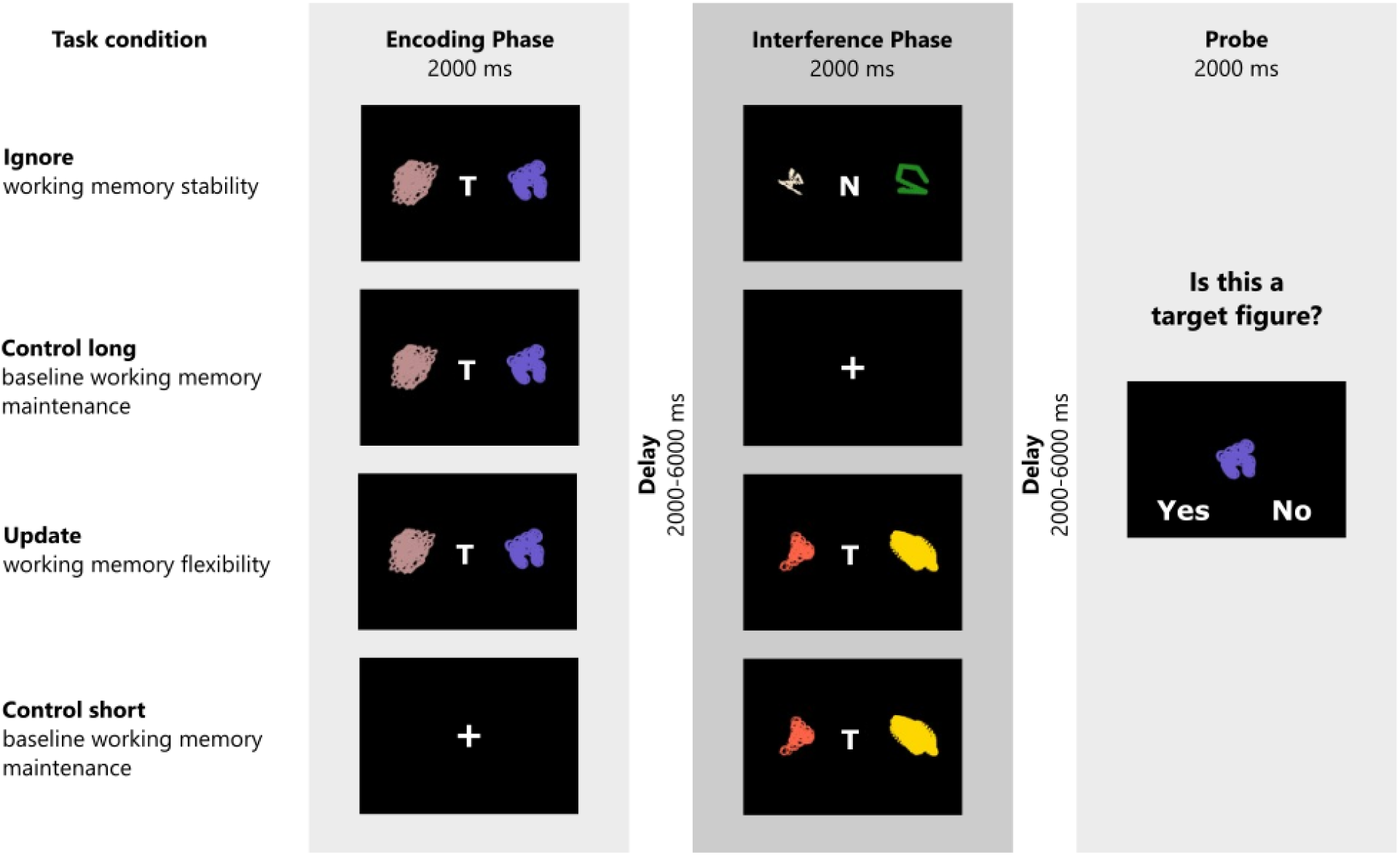
Schematic illustration of the task structure and experimental conditions. The task consists of three task phases. In the encoding phase, participants have to remember two target stimuli (signaled by the letter “T”), or are presented with a centered cross (short control trials). In the interference phase, participants either have to ignore two non-target stimuli (ignore trials; signaled by the letter “N”) or allow two new stimuli (again marked by a “T”) to replace the previously remembered target stimuli (update trials). No-interference trials (short and long control) do not require any manipulations in the interference phase. At the end of each trial, participants evaluate whether a presented figure was a target figure or not. Figure copied from Hartmann et al. (2023) with permission.

### 2.4. Genotyping

Peripheral blood samples were drawn using an EDTA monovette (2.7 ml EDTA S-Monovette, SARSTEDT AG & Co. KG, Nümbrecht, Germany). Analysis of the blood samples to extract SNP information was performed in the lab for ‘Adiposity and diabetes genetics’ at the Medical Research Center, University Leipzig, Leipzig, Germany. Genomic DNA was extracted from white blood cells with the QuickGene DNA whole blood kit S (Kubaro Industries LTD., Osaka, Japan). Genotyping of SNPs was performed with Applied Biosystems TaqMan® Genotyping Assays and the TaqMan Genotyping Mastermix on a TaqMan®7500 Real-Time PCR System (all supplied by Applied Biosystems, Foster, USA) according to the manufacturers’ protocol. For COMT Val158Met the allele frequency of the Val allele was 43.5 % and the allele frequency of the Met allele was 56.5 % (15 Val homozygotes, 37 Val/Met heterozygotes, 25 Met homozygotes). The genotype distribution for COMT Val158Met did conform to Hardy-Weinberg Equilibrium, χ2(1) = 0.003, p = .950. The allele frequency of Taq1A’s A1 allele was 27.3 % and the allele frequency of the A2 allele was 72.7 % (36 A1 carriers, 41 non-carrier). The genotype distribution for Taq1A was in Hardy-Weinberg Equilibrium, χ2(1) = .01, p = .918.

### 2.5. Statistical analyses of behavioral data

All behavioral analyses were done in R in RStudio v4.0.2 (R Core Team, 2015; RStudio Team, 2016). Generalized linear mixed models (GLM) of the *lme4* package were used to analyze the two performance measures of the task: accuracy and RT. It should be noted that the task was not speeded and therefore reaction times were not our primary outcome measure. We ran separate models for each of the performance measures. To first test our main assumption that obesity status is associated with working memory updating and maintenance, we built a model including the between-subject factor group (obese vs lean) and the within-subject factors temporal delay [short (update & ctrl short) vs long (ignore & ctrl long)] and interference [yes (ignore & update) vs no (ctrl short & ctrl long)].

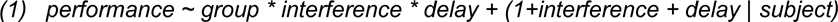

To assess group and condition effects on accuracy, we ran a logistic regression using *glmer()* with a binomial link function. We used trial-by-trial information for each subject with binary coded response (0 = incorrect; 1 = correct). Reaction time analyses were done using linear regression (*lme4::lmer*), again with trial-by-trial information for each subject. Only correct trials were included in this analysis.

For our exploratory analyses, regarding the dopaminergic gene variants, we augmented model 1 with the between-subject factors Taq1A genotype (model 2a) or COMT Val158Met (model 2b). We corrected the results for these secondary analyses for multiple comparisons by multiplying the p-value by 3.

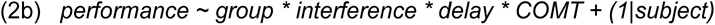

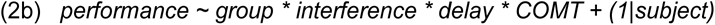

We deviate from our pre-registered analysis here. In our pre-registration, we planned to recode Taq1A and COMT polymorphism according to an equilibrium model proposed by Reuter et al. (2006). This model entails an interaction effect of these two SNPs, where striatal DRD2 density (A1-carrier vs. non-carrier) and COMT activity in the prefrontal cortex (val-carrier vs. non-carrier) are grouped according to whether they are balanced (A1+/Val+; A1-/Val-) or unbalanced (A1+/Val-; A1-/Val+). However, given that 1) each SNP alone has a very small effect size and 2) we would end up with very small gene-gene group sizes, we decided against adopting the equilibrium model and went with *post hoc* grouping of COMT and Taq1A genotypes individually. For all analyses including Taq1A, we grouped A1 homozygotes and A1/A2 heterozygotes as A1-carriers and non-carriers, respectively. We did so as the frequency of the Taq1A A1 allele is low in the general population (Noble, 2003). For all analyses including COMT Val158Met, we grouped participants into Val/Val, Val/Met, or Met/Met allele combinations. For the sake of transparency, we nevertheless ran the preregistered analyses and report the results in the supplementary materials (Table S3).

### 2.6. Functional Brain Imaging

Scans were conducted in the same manner as in Hartmann et al. (2023). A Siemens 3T Skyra magnet resonance imaging system was used. The structural sequence was a T1-weighted MP2RAGE (magnetization prepared two rapid gradient echo), 192 slices (interleaved), 1.0 x 1.0 x 1.0 mm voxel size, field of view = 256 mm, flip angles α1 = 4°, α2 = 6°, retention time = 7000 ms, inversion time 1 = 945 ms, inversion time 2 = 3770 ms. The functional scan sequence was a T2*-weighted less voids EPI (echo-planar imaging) sequence, multiband (multi-band factor 3), 60 slices (interleaved), 2.5 x 2.5 x 2.5 mm voxel size, 0.25 mm interslice gap, field of view = 204 mm, flip angle α = 80°, retention time = 2000 ms, echo time = 22 ms. Participants were scanned using a 32-channel head coil.

### 2.7. fMRI Preprocessing

All fMRI data was preprocessed in the same manner as in Hartmann et al. (2023). SPM12 (Welcome Department of Imaging Neuroscience, London, UCL, London, UK) within Matlab 9.10 (Mathworks Inc., Sherborn, MA, USA) was used. First, all functional runs were slice-time corrected (referenced to the middle slice of the functional volume), and voxel-displacement maps were computed based on the field maps. Thereafter, the images were realigned and unwarped, and segmentation (including skull-stripping) was performed. Functional scans were also co-registration to the structural T1 image, and spatially normalized to MNI (Montreal Neurological Institute) space. The normalized images were then smoothed using an 8 mm 3D FWHM Gaussian kernel. Image quality (i.e. Field-map correction, head motion, and normalization) were checked for each participant individually. No participant had to be excluded based on these data checks.

### 2.8. Imaging Data Analysis

Imaging data were missing for two participants with obesity because they were not eligible for the scanner and performed the task behaviorally only. Imaging data were analyzed using the summary statistics approach (Mumford & Nichols, 2009; Holmes & Friston, 1998). On the first level, we modeled all task events, i.e. ignore, update, fixation cross (control short) and initial target stimuli (update) during the encoding phase, fixation cross (control long), and target stimuli (short no-interference) during the interference phase, probe event, feedback screen, and participant’s response. Additionally, we computed the contrast ignore minus update (and vice versa) for each participant, which was later submitted to the second-level group analysis. We deviated from our pre-registration by using the contrast update minus ignore (or vice versa), instead of computing a contrast for each condition of interest vs. its respective control condition. At the time of pre-registration, we intended to include the control conditions in the first-level contrasts, in order to control for the difference in temporal delay until probe. However, since the actual brain activity at the time of ignoring/update is independent of delay, there is no need to control for this. Additionally, contrasting the conditions of interest with each other at the first level will control for within-subject error. We still report the results obtained from the originally planned design matrix for the sake of transparency in the supplementary materials (see Table S4). Finally, to account for potential effects of motion on our results, we also included the six motion regressors in our first-level models.

At the second level, we used a two-sample T-Test comparing the first-level ignore minus update contrast among groups. We looked at the group comparison, as well as the activation elicited by each condition. We used a region of interest (ROI) approach for the analyses of PFC and striatal ROIs. ROI selection was based on activation-based t-maps obtained from the previous study by Fallon et al. (2017, see Figure S1). To investigate the possible influence of COMT Val158Met and Taq1A, we ran two additional full factorial models including the factors genotype (for COMT: Val/Val vs Val/Met vs Met/Met; for Taq1A: A1-carrier vs non-carrier). And the factor group (obese vs. lean). We again took the first-level contrast update minus ignore. For all second level analyses, the alpha level for significant clusters was set to 0.05 (at peak-level) with small volume family-wise error correction using random field theory. The cluster-defining threshold was set to 1.

### 2.9. Brain-behavior analyses

To test whether behavioral performance on the two task conditions of interest (ignore and update) relates to condition-related BOLD signal in the striatum or PFC, we extracted mean beta values per subject from significant PFC and putamen clusters identified in our ROI analysis, and entered these as further variables of interest in separate GLMs. Because significant clusters were based on the main effect of ignore and update only, we did not include the two control conditions in these models. We hence had group (obese vs. lean) and mean beta as between-subject factor and condition (update vs. ignore) as within-subject factor in these models. We ran separate linear models for each identified ROI and corrected the results for multiple comparisons, multiplying p-values by 2 (for the 2 striatal clusters) and by 3 (for the 3 frontal clusters):

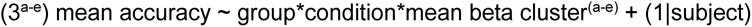

Further, in order not to miss any relevant effects that might occur beyond our regions of interest, we added mean accuracy from update minus ignore trials as an additional regressor in the second-level group design matrix and looked at its main and group effect across the whole brain. This way, we could augment our analysis to the whole brain and see whether there is a relation between condition-specific BOLD signal and behavioral performance in any brain area.

## 3. RESULTS

### 3.1. Group descriptives

Thirty-nine participants were assigned to the group with obesity (BMI: *M* = 33.70 kg/m2, *SD* = 2.77 kg/m2, min = 30.18 kg/m2, max = 41.65 kg/m2) and thirty-nine to the lean group (BMI: *M* = 22.86 kg/m2, *SD* = 1.69 kg/m2, min = 18.51 kg/m2, max = 25.12 kg/m2). The groups were matched for age (<1.5 years difference) and gender. Mean age for the group with obesity was 30.76 years (*SD* = 5.90 years, *min =* 23 years, *max =* 49.75 years). Mean age for the lean group was 30.64 years (*SD* = 5.89 years, *min =* 22.08 years, *max =* 49.25 years). There were fourteen males in each group.

There was no significant difference between groups on any of the neuropsychological tests (all p-values > 0.421). Yet, there was a trend-significant difference in IQ (p = 0.067), with lean participants having a slightly higher IQ score (mean = 109.64, SD = 9.03) than participants with obesity (mean = 105.61, SD = 10.06). Furthermore, we found group differences on some of the questionnaire scores: Participants with obesity scored significantly higher on the Eating Disorder Examination-Questionnaire, the Three-Factor-Eating-Questionnaire cognitive restraint and disinhibition subscales, the Barratt Impulsiveness motor subscale, as well as on the Food Choice Questionnaire emotion and guilt subscales (see Table S1 for a full list of questionnaire descriptives and statistical values).

### 3.2. No Evidence for Differential Working Memory Updating in Obesity

In line with previous studies (Fallon et al., 2018 & 2019), we observed a significant main effect of interference (OR = 1.13, z = 2.928, p = 0.003, Fig. 2A) and delay (OR = 0.78, z = -5.556, p < 0.001, Fig. 2B). Simple effects analyses showed that trials with a short delay had higher probability to be correct (mean = 0.94, 95%CI = [0.91 - 0.96]) than trials with a longer delay (mean = 0.91, 95%CI = [0.88 - 0.93], OR = -0.49, z = -5.55; p < 0.0001). Trials whithout interference had significantly higher probability to be correct (mean = 0.91, 95%CI = [0.88 - 0.93]) than trials with interference (mean = 0.87, 95%CI = [0.83 - 0.90], OR = 0.24, z = 2.928, p = 0.003). The interaction between interference and delay was not significant (OR = 1.01, z = 0.171, p = 0.864), indicating that ignoring did not have a significantly different effect on recall compared to updating after taking temporal differences into account.

**Figure 2.**
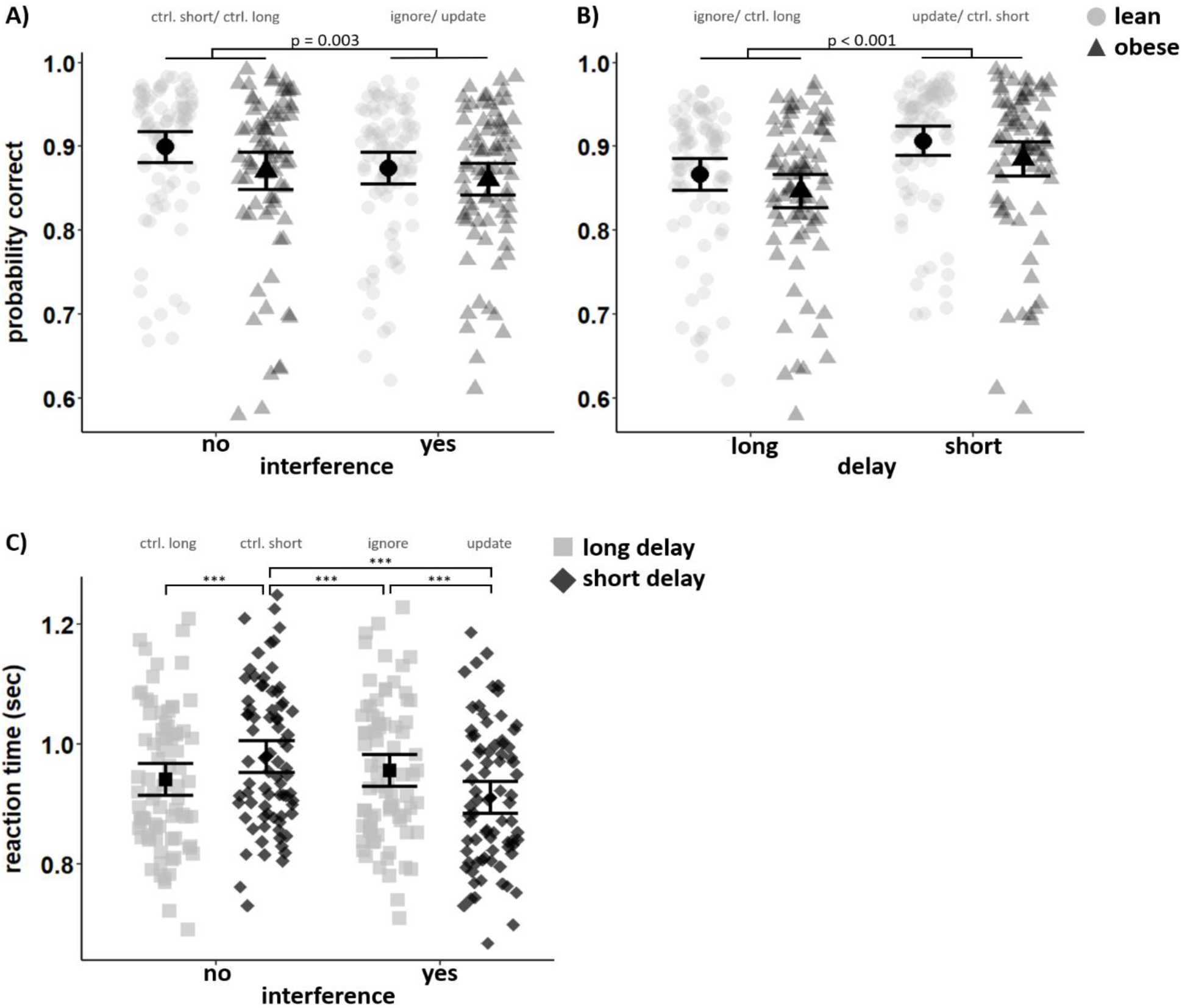
Results for model 1. There was a significant main effect of interference **(A)** and delay **(B)** on accuracy. For reaction times **(C)** there was a significant interaction effect of interference and delay. Means and standard errors are displayed.

Against our main hypothesis, however, there were no differences in condition-specific task accuracy between participants with and without obesity (group*interference*delay: OR = 1.05, 1.626, p = 0.104). There was also no significant main effect of BMI group on accuracy (OR = 1.11, z = 0.983, p = 0.326), neither did group interact with delay (OR 0.98, z = -0.409, p = 0.683) nor interference (OR = 1.03, z = 0.891, p = 0.373). Since variations in working memory performance might partly be explained by other confounding factors such as age, IQ, baseline working memory capacity (as assessed by digit span), focus and tiredness during the task, or the group differences in our questionnaires, we re-ran our analysis including those factors to control for their potential influence. Adding those variables to our main model revealed no substantial difference from our previous results (see Table S2).

Analysis of reaction time revealed a similar picture, with no effects of group on overall reaction time (β = -0.00, t(1) = -0.02, p = .996), or any two-or three-way interactions of group and interference and delay (all p-values > 0.365). However, there was a main effect of interference (β = 0.01, t(1) = 5.3, p < 0.001). Moreover, we observed a significant interaction between interference and delay (β = -0.02, t(1) = -8.29, p < 0.001, Fig. 2C). Reaction times were fastest in the update condition (M = 0.911sec, SD = 0.0145sec), followed by long control trials (M = 0.941sec, SD = 0.0145sec), then by ignore trials (M = 0.956 sec, SD = 0.0145sec), and finally by short control trials (M = 0.979sec, SD = 0.0145sec). All *post hoc* comparisons were significant (all corrected p < 0.006), except the one for ignore vs. control_long (p = 0.157).

#### 3.2.1. Taq1A Genotype Modulates Group-Dependent Task Performance

On a more exploratory basis, we further examined whether the genetically determined availability of dopamine in the PFC (COMT Val158Met) or striatal D2 receptor density (Taq1A) would modulate working memory maintenance or updating. Chi-square tests revealed no BMI group differences in the distribution of COMT Val158Met, χ2(2) = 1.29, p = .523, and Taq1A genotypes, χ2(2) = .739, p = .691.

With respect to accuracy, we found a significant interaction between group and Taq1A (OR = 0.79, z = -2.415, pcorrected = 0.032, Figure 3A). All other main or interaction effects of Taq1A were non-significant (all corrected p-values > 0.1). In line with the previous analysis, the main effect of delay and interference remained significant (corrected pdelay < 0.001, corrected pinterference = 0.01). Simple effect analysis revealed that the interaction between group and Taq1A was mainly driven by a group difference in A1-carriers, such that lean A1+ tend to have higher probability to score correct (mean = 0.91, 95%CI = [0.89 - 0.95]) than A1+ in the obese group (mean = 0.87, 95%CI = [0.80 - 0.92], OR = 0.644, z = 2.269, p = 0.023). There was no such effect visible for non-carriers of the A1 allele (OR = 0.87, z = -1.095, p = 0.273), nor was there a difference in whether or not one carries the A1 allele in the group with obesity (OR = 1.27, z = 1.723, p = 0.085). Although the four-way interaction of group, genotype, delay, and interference did not reach significance, we were wondering whether the difference in the A1+ genotype group followed a consistent pattern across all four conditions. Therefore, we built a subset of data including A1+ individuals only and ran group comparisons for each condition separately. The results were corrected for multiple comparisons by multiplying the p-value by four, as we ran four additional analyses. Results show that there was a group difference in the update condition only (pcorrected = 0.039), but not so in the other conditions (all corrected p > 0.281). See Figure 3B. With respect to reaction times, we found no significant interaction between Taq1A, group, interference and/or delay (all corrected p-values >0.117). The interaction between interference and delay (compare model 1) stayed significant (p < 0.001).

**Figure 3.**
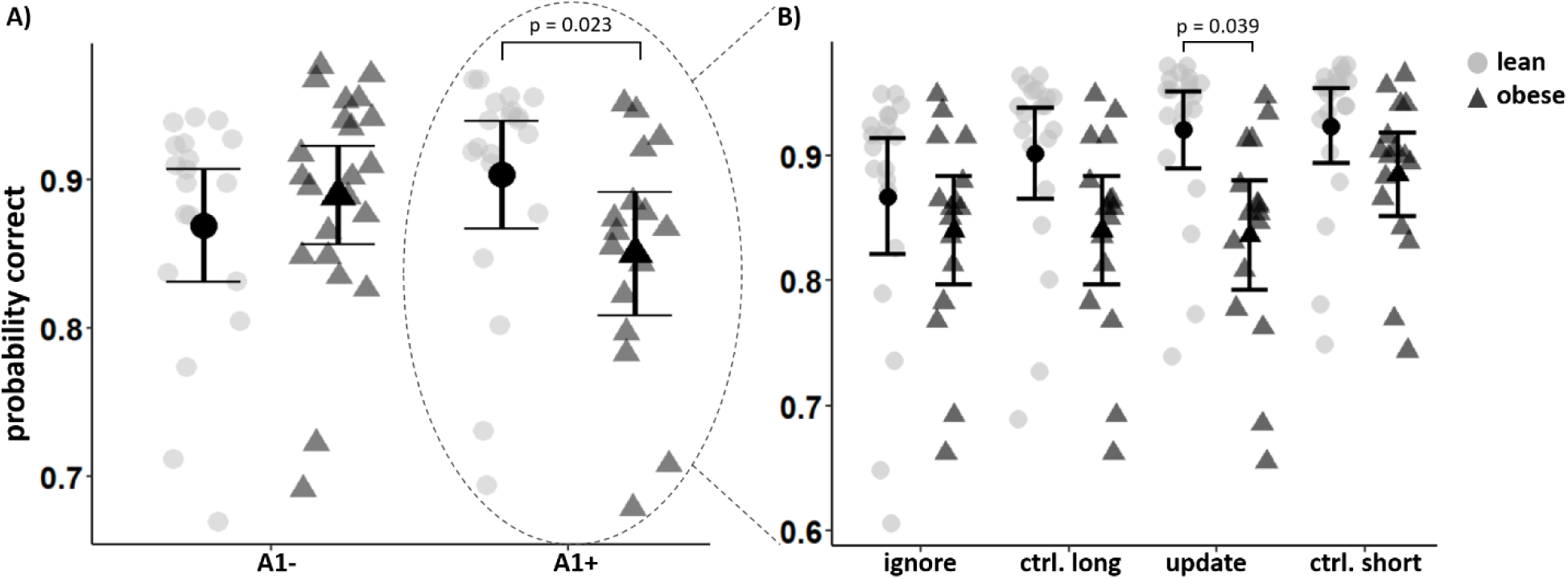
Results from Model 2a. **A)** There was a significant interaction between Taq1A genotype and group on overall performance. Post hoc analyses showed that A1-carrier in the lean group perform better than A1-carriers in the group with obesity. **B)** Follow-up analyses on the A1-carriers only. There was a difference in performance only in the update condition, but not so in the other conditions. Means and standard errors are displayed.

When looking at the effect of COMT genotype (model 2b) on accuracy, there were no significant main- or interaction effects (all corrected p-values > 0.18). Similarly, there was no effect of COMT on reaction time (all corrected p-values > 0.168). The interaction between interference and delay remained significant, however (pcorrected= 0.011).

### 3.3. fMRI

#### 3.3.1. No Group Differences in Neural Activation Related to Ignoring and Updating

Against our main hypotheses, we did not observe significant differences in condition-specific BOLD activity between groups. We do find the expected significant main effects of task conditions, however. Similar to previous reports, BOLD signal changes in middle and superior PFC as well as in parietal and occipital gyri, when contrasting ignore versus update (Fig. 4, upper panel). Likewise consistent with previous reports, the reverse contrast – update relative to ignore – produced significantly increased BOLD signal in the left and right dorsal striatum and the right thalamus as well as in occipital gyri (Fig. 4, lower panel). A full list of significant condition-specific clusters is presented in Table 1.

**Figure 4.**
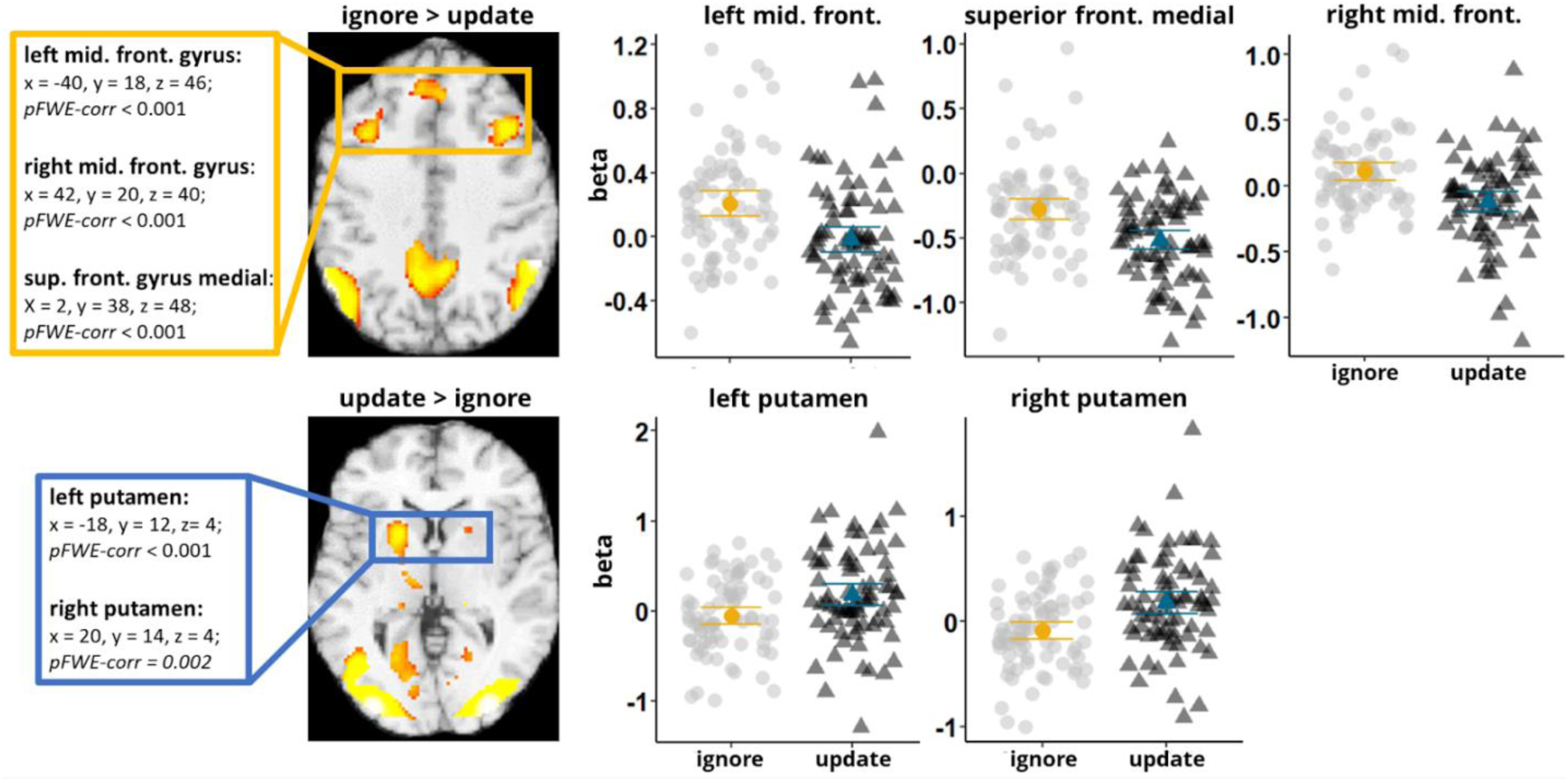
Bold signal related to conditions of interest. **Upper panel:** Ignore minus update elicited BOLD response in three frontal regions (yellow square) as well as in parietal and occipital gyri. Mean betas from the frontal regions were higher in the ignore as compared to the update condition. Median and standard errors are displayed. **Lower panel:** Update minus ignore elicited BOLD responses in the left and right putamen (blue square), as well as in the right thalamus and occipital regions. Mean betas in the left and right putamen were higher in the update condition as compared to the ignore condition. Median and standard errors are displayed

**Table 1.** Overview of all clusters with significant neural activation for updating and ignoring.

| contrast | brain region | cluster extent | p (FEW-corr) | T | MNI coordinates |  |  |
| --- | --- | --- | --- | --- | --- | --- | --- |
|  |  |  |  |  | x | y | z |
| ignore > update | left angular gyrus | 1424 | 0.00 | 9.28 | -54 | -58 | 36 |
|  |  |  |  | 6.57 | -42 | -68 | 48 |
|  | right supramarginal gyrus | 716 | 0.00 | 8.94 | 58 | -46 | 40 |
|  |  |  |  | 8.03 | 50 | -52 | 26 |
|  | middle temporal gyrus | 285 | 0.00 | 7.94 | -58 | -26 | -10 |
|  | precuneus | 892 | 0.00 | 7.72 | -8 | -54 | 42 |
|  |  |  |  | 7.36 | 8 | -54 | 40 |
|  | **left middle frontal gyrus** | **336** | **0.00** | **7.56** | **-40** | **18** | **46** |
|  | **superior medial frontal gyrus** | **403** | **0.00** | **6.65** | **2** | **38** | **48** |
|  |  |  |  | 5.81 | -4 | 44 | 30 |
|  |  |  |  | 5.64 | -4 | 42 | 38 |
|  | left middle temporal gyrus | 26 | 0.01 | 6.59 | -60 | -44 | -2 |
|  | **right middle frontal gyrus** | **265** | **0.00** | **6.52** | **42** | **20** | **40** |
|  |  | 149 | 0.00 | 6.52 | 32 | 56 | 6 |
|  | right middle temporal gyrus | 49 | 0.00 | 6.33 | 64 | -28 | -8 |
|  | left superior frontal gyrus | 80 | 0.00 | 5.61 | -24 | 56 | 16 |
|  | right parietal operculum | 2 | 0.03 | 4.98 | 56 | -26 | 22 |
|  | right orbital inferior frontal gyrus | 1 | 0.04 | 4.93 | 48 | 30 | -10 |
|  | right central operculum | 1 | 0.04 | 4.89 | 54 | -20 | 18 |
| update > ignore | left occipital lobe | 6477 | 0.00 | 16.18 | -40 | -72 | -8 |
|  |  |  |  | 15.67 | -30 | -58 | -16 |
|  |  |  |  | 14.92 | -36 | -84 | -6 |
|  | right occipital lobe | 4898 | 0.00 | 15.43 | 28 | -88 | -6 |
|  |  |  |  | 14.96 | 36 | -64 | -12 |
|  |  |  |  | 14.27 | 36 | -84 | -2 |
|  | supplementary motor cortex | 589 | 0.00 | 13.08 | -4 | 6 | 64 |

**Table 1.** continued
|  |  |  |  |  |  |  |  |
| --- | --- | --- | --- | --- | --- | --- | --- |
| update > ignore |  |  |  | 7.09 | -6 | 16 | 42 |
|  | left inferior frontal gyrus | 1204 | 0.00 | 11.61 | -44 | 6 | 26 |
|  |  |  |  | 10.73 | -48 | 0 | 50 |
|  | right inferior frontal gyrus | 459 | 0.00 | 9.23 | 46 | 6 | 26 |
|  | left thalamus | 69 | 0.00 | 9.17 | -22 | -30 | -2 |
|  | right thalamus | 39 | 0.00 | 8.95 | 20 | -30 | 2 |
|  | **left putamen** | **626** | **0.00** | **8.12** | **-18** | **12** | **4** |
|  |  |  |  | 7.96 | -12 | -14 | 6 |
|  |  |  |  | 7.33 | -18 | 2 | 8 |
|  | right middle frontal gyrus | 63 | 0.00 | 7.54 | 50 | 42 | 12 |
|  | brain stem | 20 | 0.01 | 6.97 | 8 | -30 | -4 |
|  |  | 18 | 0.01 | 6.7 | -6 | -30 | -6 |
|  | right middle frontal gyrus | 42 | 0 | 5.85 | 26 | -4 | 48 |
|  | cerebellum | 5 | 0.03 | 5.69 | -6 | -70 | -14 |
|  | **right putamen** | **13** | **0.01** | **5.39** | **20** | **14** | **4** |
|  | left middle frontal gyrus | 14 | 0.01 | 5.31 | -48 | 36 | 16 |
|  | right orbital gyrus | 1 | 0.04 | 5.18 | 22 | 34 | -14 |
|  | right orbital gyrus | 1 | 0.04 | 5.18 | 22 | 34 | -14 |

Note: bold clusters represent the clusters shown in Figure 4

#### 3.3.2. Genotypes do not interact with condition-specific BOLD activity

Furthermore, we aimed to investigate group differences in condition-specific BOLD activity between Taq1A and COMT Val158Met genotypes. Against our expectations, neither COMT Val158Met nor Taq1A genotype revealed significant voxels nor did we find any interactions with group or condition.

### 3.4. Brain-behavior correlations

To test whether behavioral performance on our two task conditions of interest relates to previously identified striatal and prefrontal ROI activity (see Figure 4), we next regressed mean betas obtained from these clusters onto mean accuracy of each task condition (model 3a-e).

Results for the analysis with striatal ROIs showed a significant interaction between condition and beta values in the right putamen (β= 0.05, t(1) = 2.775, pcorrected = 0.012), and a significant interaction between condition and beta values in the left putamen (β = 0.04, t(1) = 2.307, pcorrected = 0.042). Higher beta values relate positively to performance in the ignore condition, but negatively to performance in updating (Fig. 5). All other two-way interactions and the three-way interaction between group, condition and betas were non-significant (all corrected p > 0.431). When examining the effects of frontal ROI activity, we found no significant interaction between mean beta, group and condition (all corrected p-values > 0.15).

**Figure 5.**
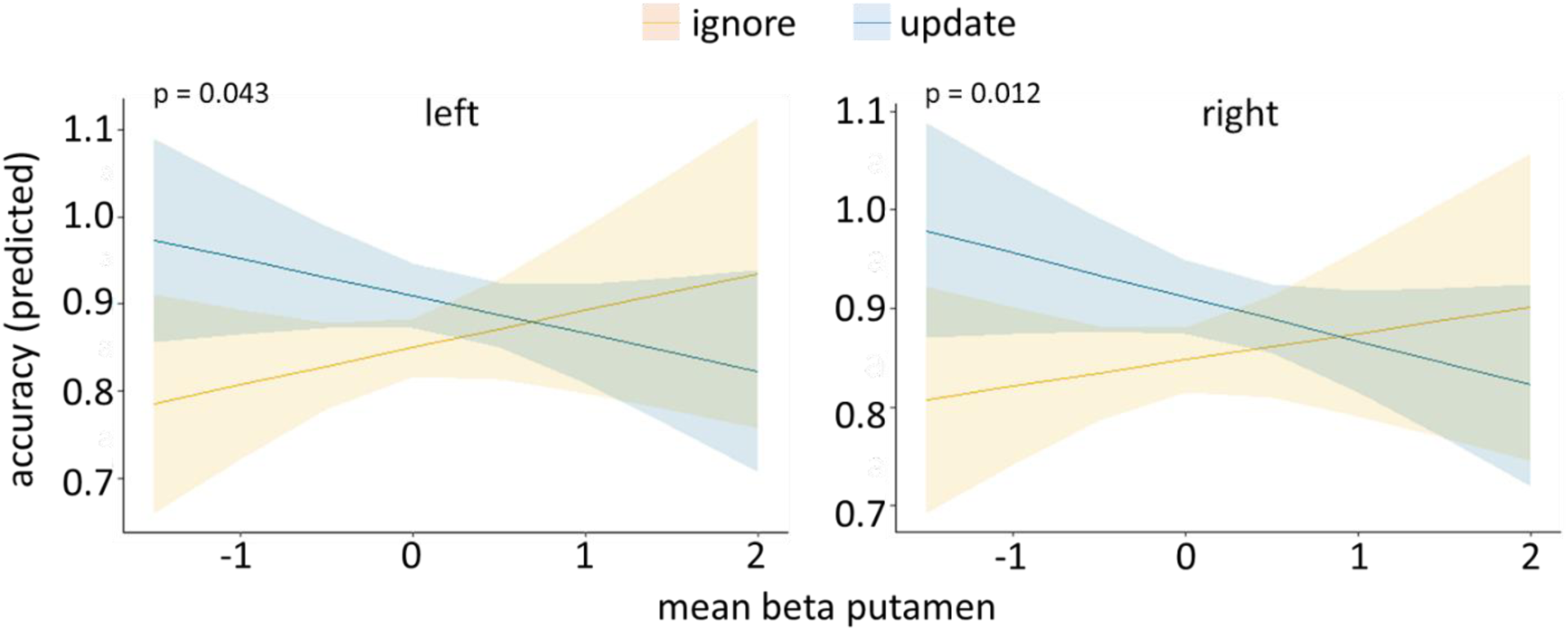
Relationship of accuracy and mean beta values from left and right putamen ROI. Shaded areas represent the 95% confidence level interval.

Next, the above analysis was corroborated by adding condition-specific mean accuracy (update minus ignore) as an additional regressor in our second-level group comparison. This way, we could see whether performance is related to activity in any other brain region. Results did not yield any significant voxels.

## 4. DISCUSSION

In this study, we aimed to investigate whether individuals with obesity differ from lean in terms of working memory maintenance and/or updating. Contrary to our main hypothesis, we could not find evidence for group differences on these two working memory sub-processes, neither on the behavioral level (accuracy and reaction time) nor on the neural level. Still, we did observe the expected brain activations elicited by ignoring or updating, however. Interestingly, we could observe a group-dependent effect of Taq1a genotype on overall task performance, which was not reflected in comparable differences in brain activation though.

The lack of group differences in overall working memory performance was surprising, as there is numerous evidence pointing toward diminished working memory functioning in obesity (Yang et al., 2018, 2019, 2020; Gonzales et al., 2010; Coppin et al., 2014). Additionally, in a recent study done in our lab (Hartmann et al., 2023) we found evidence of a link between BMI and performance on the current task: higher BMI was related to worse overall performance. These discrepancies might be due to the fact that in our previous study, BMI did not include the range for obesity (BMI > 30kg/m²) and was a continuous, rather than a grouping variable.

Although we did expect to find working memory differences in our groups, a closer look at the literature reveals that the evidence with respect to an association between obesity and working memory is mixed. While there are several studies reporting worse working memory in obesity, there are also others that did not observe such an effect (Calvo et al., 2014, Schiff et al., 2016, Alarcon et al., 2016). Additionally, group differences often become apparent only in combination with mediating factors, such as insulin resistance, inflammation, or mood (Gonzales et al., 2010; Yang et al., 2020; Fitzpatrick et al., 2013). Although mood did not seem to affect our findings (see control analyses, Table S2, i.e. BDI), our study did not include any measures for inflammation or insulin resistance. We therefore cannot exclude the possibility that (potentially subtle) group differences in working memory are masked by those effects. Moreover, it appears that the task used might influence whether or not one can detect working memory deficits in obesity. Those studies that did not find group differences, employed tasks whose outcomes were binary (Calvo et al., 2014, Schiff et al., 2016) or tasks that had a small memory load (Alarcon et al., 2016). Our task also had a binary outcome and a small memory load. At the same time, participants generally performed very well (overall median % correct = 90.6), which concurs with the performance reported in those studies with null-findings. This leads us to speculate that our task might not have been sensitive enough to capture the potentially small differences in working memory among our groups. Future studies that aim to probe working memory in obesity should take this into consideration and try to boost explanatory variance by i) experimentally accounting for potential mediating factors, and ii) using more sensitive tasks, employing continuous outcome measures, and higher WM loads.

Another possible account for the mixed evidence with respect to working memory deficits in obesity might stem from the fact that most studies do not clearly disentangle the two specific working memory sub-processes (distractor-resistance vs. update). In our study, we aimed to address this gap, by specifically probing maintenance and updating in obesity. However, we could not find evidence of alterations in either of the two processes. This, on the one hand, could be due to the task - as mentioned above. On the other hand, we speculate that the putative dopaminergic differences among our BMI groups might not have been pronounced enough to reveal distinct effects of condition. It’s well-established that alterations in working memory gating are contingent on baseline dopaminergic tone (e.g. Furman et al., 2021; Cools & D’Esposito, 2011; Cools & Robbins, 2004). This relationship typically follows an inverted-U-shape with respect to dopamine: Subjects with a low baseline tend to show improvement in maintenance when dopamine levels are elevated, whereas those with a high baseline may experience performance decline (Fallon et al., 2019; also see Cools & Robbins, 2004 for an extensive review). Interestingly, these alterations tend to manifest as a trade-off between maintenance and updating, i.e., as one improves, the other tends to deteriorate. While a growing body of evidence links obesity to alterations in dopamine transmission (Horstmann et al., 2015b), the exact nature of this relationship remains under debate and appears to be highly complex (Janssen and Horstmann, 2022). Considering the likelihood of the inverted-U-shaped influence of dopamine, it’s conceivable that the groups in our study could have shared similar starting points on this dopaminergic parabola, which in turn could account for the absence of significant differences in maintenance or updating.

Previous studies, which were successful in showing dopamine-dependent effects using the same working memory task, employed drug manipulations (i.e. within-subject designs), and/or continuous outcome measures (Fallon et al., 2017, Fallon et al., 2018, Fallon et al., 2019). These studies thus had a higher statistical power, handling stronger effects. This emphasizes that strong variations in dopamine levels may be needed to uncover disparities in the ability to maintain and update working memory. In support of this notion, also Fallon and colleagues (2017) could not observe between-but only within-subjects differences in WM gating. Further, in a previous study (Hartmann et al., 2023), we could also not find between-subjects differences in working memory gating among two diet groups that are presumably comparable to differ in terms of dopamine signal transmission. With the help of more direct measures of endogenous dopamine levels, such as PET, future studies will be able to shed more light on the relationship of obesity and dopamine-dependent cognition.

In conclusion, it seems that our null findings might be explainable by the combination of two factors: insufficient sensitivity of the task, on the one hand, and too subtle dopamine/working memory differences among groups, on the other.

### Group-genotype Interaction on overall task performance

In line with the speculation that the obesity-associated dopaminergic differences might be too subtle in our sample, we see that group effects do emerge when we account for previously unexplained dopamine-related variance in the outcome variable (i.e. Taq1A). We found a significant interaction effect of group and Taq1A genotype on overall performance, such that obese A1-carriers performed worse than lean A1-carriers.

Our findings resemble the obesity x Taq1A interaction pattern found by Ariza et al. (2012). Just like us, Ariza and colleagues show that in obese participants, carrying the A-allele leads to worse performance on a working memory task. Furthermore, evidence from Persson et al. (2015) and Xin et al. (2018) shows that the effects of the A-allele on cognition become apparent in older adults only, a population whose dopamine system supposedly is compromised. We likewise speculate that our findings can be explained by a potentially compromised dopamine system in obesity (Horstmann et al., 2015).

Critically, carrying the A-allele has been linked to reduced striatal D2 receptor density (Thompson et al., 1997; Pohjalainen et al., 1998; Jönsson et al., 1999; Eisenstein et al., 2016) and is often considered a risk factor for obesity (Cardel et al., 2019, Kibitov et al., 2020, Johnson & Kenny, 2010) or other addictive-like behaviors (Smith et al., 2008; Nacak et al., 2012; Comings & Blum, 2000, Gorwood et al., 2012). Of course, we cannot say what, if any, came first - dopamine alterations or obesity. But it could very well be that obesity-promoting behavior (diet and/or inactivity) led to dopamine changes, which then affect those with the “vulnerable” genotype more strongly. Both our and previous findings are thus in line with the resource-modulation hypothesis (Lindenberger et al., 2008), proposing that genetic effects on cognition are larger when resources (such as dopamine in obesity) are limited.

The impact of Taq1A genotype was not condition-dependent, however. Obese A1 carriers consistently performed worse on all four conditions. Yet, this difference was most pronounced in the update condition. This concurs with the fact that the A1 allele has been linked to differential performance on tasks that require updating of working memory representations (Stelzel et al., 2010, Persson et al., 2015; Li et al. 2019). However, the sample sizes of these studies were larger, or they had continuous predictor variables as opposed to our categorical one (group). Consequently, those studies were better powered to identify potentially small condition-genotype interaction effects. As our genotype analysis was of a more exploratory nature, it might be that it was not sufficiently powered to reveal those higher-order interaction effects. The association of obesity, genotype, and dopamine-dependent cognition needs to be investigated further in larger samples to verify our preliminary and explorative findings.

### Imaging

We observed the expected BOLD activation for the respective condition. We can hence replicate previously observed activation patterns of WM maintenance and updating (see Fallon et al., 2017, Hartmann et al., 2023). However, against our hypothesis, we were not able to find evidence for group differences in BOLD activation related to these two processes. In this respect, our imaging results are consistent with our behavioral results, indicating that our sample with obesity did not have differential working memory updating and/or maintenance. Interestingly, the activation pattern during ignoring largely resembles the activity of the default mode network (DMN), a network associated with off-task mind wandering (Raichle et al. 2001, 2015). It could hence be that trying to ignore distractors can be equated with retracting from the task at hand (i.e. getting into an off-task mode) to guard the already encoded items, making ignoring seem like a more passive task.

Furthermore, our behavioral findings on Taq1A genotype-dependent group differences were not reflected on the level of the brain. Previous studies, that were able to detect Taq1A-related differences in striatal BOLD activation in paradigms probing cognitive updating, had larger sample sizes (Persson et al., 2015), or less complex study designs and male samples only (Jocham et al., 2009).

### Activation in the putamen affects condition-dependent performance

Our results indicate that activation of the putamen in response to updating was behaviorally relevant, but that - in keeping with our earlier results - this was not differential for groups. We found an interaction effect of beta values in the putamen and condition on task performance. Higher beta values were associated with numerically better performance on ignore but with worse performance on update. We postulate that this is a result of go/no-go path activation in the striatum. High striatal activation would hence represent inhibitory no-go path activation, promoting more efficient ignoring, while less activation would represent go-path activation, resulting in better updating. At the system level, most models speculate that opening and closing the gate to WM should be a collaborative action between the basal ganglia and frontal cortex. Our brain-behavior relationships might therefore reflect DLPFC top-down modulation. In the ignore condition, the more the DLPFC suppresses striatal activity the easier it is to disregard. In contrast, in the update condition, more DLPFC-induced suppression leads to worse performance due to impaired updating. Results of an exploratory psychophysiological interaction (PPI) analysis (see supplements) support this speculation in the sense that we observed functional coupling of the the putamen with thalamus and frontal lobe - areas associated with basal ganglia go/no-go pathways (Gurney, Prescott, & Redgrave, 2001) - both, during ignoring and updating. These interactions were not behaviorally relevant, however.

### Overall conclusion

Our study seems to point towards no alterations in working memory updating or maintenance in individuals with obesity within our sample. However, our data suggest a potential role for the Taq1A genotype, particularly the A allele, in influencing this observation, hinting at a nuanced interplay between obesity, working memory gating, and this specific genotype. However, due to the limited power of the genetic analysis in our study, these findings remain suggestive and should be further investigated with larger sample sizes.

## Acknowledgements

The authors thank Susan Prejawa for assistance in study organization and financial management. Furthermore, we thank Miriam Huml, Eva Burmeister and Lisa Okhof for helping with recruitment and testing of participants. Special thanks to Prof. Arno Villringer and the entire medical-technical staff of the neuroimaging facilities at the Max Planck Institute for Human and Cognitive Brain Sciences, Leipzig. Last but not least, we thank Peter Kovac and his lab at the University of Leipzig for the analyses of our blood samples.

## Author contribution statement

NH, LD, LJ, and AH designed the study. NH and MW collected the data. NH analyzed the data. NH drafted the manuscript. HH, LJ, MW, LD, SF, and AH critically revised and approved the final manuscript.

## Data availability statement

Data will be made available upon request.

## Declaration of Generative AI and AI-assisted Technologies in the writing process

During the preparation of this work, the author used ChatGPT in order to improve readability and language. After using this tool, the author reviewed and edited the content as needed and takes full responsibility for the content of the publication

