## Supplemental Material for "Working Memory Gating in Obesity: Insights from a Case-Control fMRI Study"

**Table S1**
*Demographics, Neuropsychological Tasks, and Questionnaire Statistics*

| | | | obese | lean | statistics | | |
|---|---|---|---|---|---|---|---|
| | | | mean (sd) | mean (sd) | t-value | df | p-value |
| Demographics | Age | | 30.76 (5.90) | 30.60 (5.78) | 1.312 | 78 | 0.903 |
| | N male | | 14 | 14 | | | |
| | Years of education | | 17.38 (4.84) | 17.51 (2.75) | 1.215 | 78 | 0.886 |
| | **BMI** | | **33.70 (2.77)** | **22.87 (1.70)** | **-20.242** | **78** | **< 0.001** |
| Neuropsychological Tasks | IQ | | 105.61 (10.06) | 109.64 (9.03) | -1.514 | 78 | 0.067 |
| | TMT A (sec) | | 20.28 (4.59) | 19.84 (4.65) | 0.467 | 78 | 0.679 |
| | TMT B (sec) | | 40.01 (14.04) | 41.95 (10.39) | -0.014 | 78 | 0.494 |
| | DS forward (level) | | 6.41 (1.18) | 6.64 (1.32) | -0.201 | 78 | 0.421 |
| | DS backward (level) | | 5.23 (1.40) | 5.33 (1.26) | 0.632 | 78 | 0.735 |
| | DSST (N corr. symbol) | | 83 (12.85) | 83.38 (11.41) | 1.232 | 78 | 0.889 |
| Questionnaires | **EDEQ** | total | **1.40 (0.81)** | **0.66 (0.56)** | **-4.355** | **75** | **<0.001** |
| | **BDI** | total | **6.61 (5.21)** | **3.58 (3.95)** | **-2.622** | **77** | **0.005** |
| | BIS | anxiety | 13.64 (3.21) | 14.58 (2.87) | -0.915 | 77 | 0.182 |
| | | fear | 5.02 (1.33) | 5.37 (2.36) | -0.623 | 77 | 0.267 |
| | BAS | drive | 11.67 (2.16) | 11.63 (2.36) | 1.625 | 77 | 0.946 |
| | | reward | 15.92 (2.28) | 16.66 (1.98) | -1.111 | 77 | 0.135 |
| | | fun seeking | 11.49 (1.68) | 11.53 (2.12) | 1.482 | 77 | 0.929 |
| | BIS15 | planning | 10.13 (2.92) | 10.53 (3.05) | 1.152 | 77 | 0.560 |
| | | **motor** | **11.23 (2.71)** | **9.92 (2.64)** | **-1.831** | **77** | **0.035** |
| | | attention | 9.38 (2.79) | 9.87 (3.02) | -0.082 | 77 | 0.468 |
| | | total | 30.74 (5.91) | 30.32 (7.22) | 0.764 | 77 | 0.776 |
| | UPPS | urgency | 26.21 (5.42) | 24.95 (5.51) | -0.478 | 76 | 0.317 |
| | | premeditation | 21.55 (3.45) | 22.21 (4.51) | -0.058 | 76 | 0.477 |
| | | perseverance | 18.71 (4.54) | 18.76 (5.51) | 1.823 | 76 | 0.964 |
| | | sensation seeking | 32.58 (7.78) | 32.87 (7.78) | 1.143 | 76 | 0.872 |
| | mYFAS2 | | 0.44 (1.23) | 0.13 (0.41) | -1.033 | 77 | 0.152 |
| | FCQ | plan | 7.33 (2.99) | 6.34 (2.64) | -1.147 | 77 | 0.128 |
| | | positive reactions | 12.62 (4.26) | 11.71 (3.95) | -0.422 | 77 | 0.337 |
| | | negative reaction | 6.49 (2.35) | 6.00 (2.71) | -0.250 | 77 | 0.402 |
| | | control | 10.31 (3.86) | 8.76 (3.66) | -1.449 | 77 | 0.076 |
| | | thoughts | 11.62 (5.02) | 10.97 (4.55) | 0.149 | 77 | 0.559 |
| | | hunger | 11.36 (3.47) | 10.11 (3.85) | -1.101 | 77 | 0.137 |
| | | emotion | **9.59 (3.54)** | **7.76 (4.05)** | **-1.795** | **77** | **0.038** |
| | | environment | 12.10 (3.80) | 11.42 (3.73) | -0.177 | 77 | 0.430 |
| | | guilt | **6.28 (2.92)** | **4.55 (1.76)** | **-2.889** | **77** | **0.003** |
| | | total | 89.85 (25.39) | 79.50 (25.69) | -1.422 | 77 | 0.080 |

**Table S1**
*Demographics, Neuropsychological Tasks, and Questionnaire Statistics*

| | | obese | lean | statistics | | |
|---|---|---|---|---|---|---|
| | | mean (sd) | mean (sd) | t-value | df | p-value |
| TFEQ | restrain | **8.85 (5.03)** | **5.87 (4.53)** | **-2.403** | **73** | **0.009** |
| | disinhibition | **6.20 (2.94)** | **4.34 (2.60)** | **-2.604** | **73** | **0.005** |
| | hunger | 4.97 (2.73) | 4.55 (2.32) | -0.047 | 73 | 0.481 |
| DFSQ | | 54.87 (11.84) | 55.74 (11.77) | 0.674 | 77 | 0.749 |
| STAI | | 37.56 (8.68) | 35.92 (9.36) | -0.185 | 77 | 0.427 |
| NEOFFI | neuroticism | 8.38 (3.56) | 9.50 (3.38) | -0.988 | 77 | 0.163 |
| | extraversion | 14.26 (3.39) | 14.74 (3.44) | 0.097 | 77 | 0.539 |
| | openness | 15.00 (4.12) | 15.71 (4.72) | -0.041 | 77 | 0.484 |
| | agreeableness | 17.31 (3.87) | 17.92 (4.15) | 0.010 | 77 | 0.504 |
| | consciousness | 18.59 (3.88) | 18.37 (4.14) | 0.880 | 77 | 0.809 |

Note: TMT = Trailmaking Test (part A + B, Reitan, 1955); DS = Digit Span Test (Wechsler, 2008); DSST = Digit Symbol Substitution Test (Wechsler, 2008). EDEQ = Eating Disorder Examination Questionnaire (Hilbert et al., 2007, 2012). BDI = Beck Depression Inventory (Beck et al., 1996). STAI = State-Trait Anxiety Inventory (Spielberger et al.,1971; Laux et al., 1981). BIS15 = Barratt Impulsiveness Scale (Meule, Vögele, & Kübler, 2011). UPPS = Urgency, Premeditation, Perseverance, Sensation Seeking Impulsive Behavior Scale (Schmidt et al., 2008). BIS/BAS = Behavioral Inhibition and Behavioral Activation System Scales (Carver & White, 1994; Strobel et al., 2006). mYFAS2= modified Yale Food Addiction Scale 2.0 (Schulte & Gearhardt, 2017), FCQ-T = Food Craving Questionnaire Trait (Cepeda-Benito et al., 2000), TFEQ = Three-Factor Eating Questionnaire (Pudel & Westhöfer, 1989; Stunkard & Messick, 1985). DFSQ = Dietary Fat and Free Sugar Questionnaire (Francis & Stevenson, 2013; Fromm & Horstmann, 2019). NEOFFI = NEO personality inventory (Costa & McCrae, 2008; Körner et al., 2008).

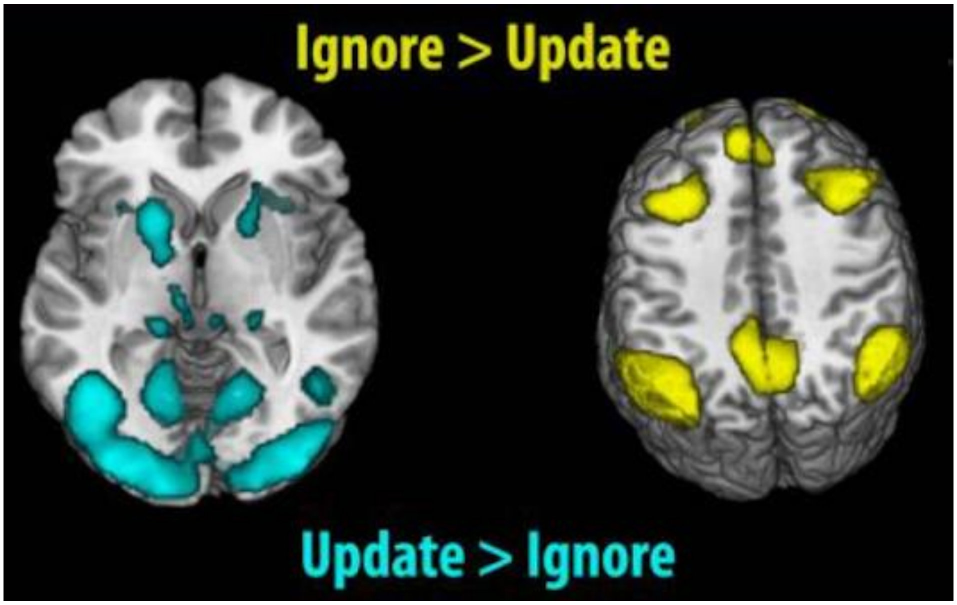


**Figure S1.** Activation-based t-maps used for our ROI analysis (see imaging data analysis). The Figure is from Fallon et al. (2017).

**Table S2**
*Full output for the model including possible cofounds*

| | accuracy | | | | |
|---|---|---|---|---|---|
| Predictors | Odds Ratios | Std. Error | CI | z-value | p |
| Intercept | 10.67 | 0.428 | 2.93 – 22.23 | 5.528 | <0.001 |
| Group | 1.07 | 0.103 | 0.86 – 1.27 | 0.625 | 0.532 |
| **Interference** | **1.09** | **0.033** | **1.03 – 1.17** | **2.735** | **0.006** |
| **Delay** | **0.84** | **0.033** | **0.79 – 0.90** | **-5.217** | **<0.001** |
| Digit span | 1.05 | 0.066 | 0.92 – 1.19 | 0.704 | 0.481 |
| **Tiredness (z-scored)** | **0.74** | **0.099** | **0.61 – 0.90** | **-3.061** | **0.002** |
| **Concentration (z-scored)** | **1.32** | **0.096** | **1.12 – 1.63** | **2.920** | **0.003** |
| Age (z-scored) | 1.09 | 0.081 | 0.92 – 1.28 | 1.003 | 0.316 |
| IQ (z-scored) | 0.92 | 0.098 | 0.76 – 1.13 | -0.881 | 0.378 |
| BDI | 0.99 | 0.019 | 0.96 – 1.03 | -0.380 | 0.703 |
| EDEQ total | 0.88 | 0.195 | 0.61 – 1.32 | -0.643 | 0.520 |
| BIS motor | 0.98 | 0.019 | 0.95 – 1.09 | -0.888 | 0.374 |
| FEV disinhibition | 0.95 | 0.040 | 0.87 – 1.02 | -1.351 | 0.177 |
| FEV restraint | 0.98 | 0.021 | 0.94 – 1.02 | -0.900 | 0.368 |
| FCQ guilt | 0.98 | 0.050 | 0.89 – 1.08 | -0.441 | 0.659 |
| FCQ emotion | 1.05 | 0.030 | 0.99 – 1.12 | 1.636 | 0.102 |
| Group * Interference | 1.03 | 0.033 | 0.96 – 1.10 | 0.841 | 0.400 |
| Group * Delay | 0.97 | 0.033 | 0.91 – 1.04 | -0.854 | 0.393 |
| Interference * Delay | 1.01 | 0.033 | 0.94 – 1.07 | 0.166 | 0.868 |
| Group * Interference * Delay | 1.06 | 0.033 | 0.99 – 1.13 | 1.735 | 0.083 |
| Observations | 9970 | | | | |
| Marginal R2 / Conditional R2 | 0.073 / 0.175 | | | | |

**Behavioral data analysis with gene-equilibrium model**

To follow our original analysis plan, we extended our main model (model 1) with the factor balance (unbalanced vs. balanced). The model hence was:

*performance ~ group * interference * delay * balance + (1|subject)*

Results show no significant interactions (all p > 0.069). The main factor of delay and interference remain significant (ps < 0.004).

**Table S3**

*Full output for the model including balanced vs. unbalanced genotypes*

| | accuracy | | | | |
|---|---|---|---|---|---|
| Predictors | Odds | Std. Error | CI | z-value | p |
| Intercept | 9.04 | 0.096 | 7.48 – 10.93 | 22.699 | <0.001 |
| Group | 1.10 | 0.096 | 0.91 – 1.32 | 0.943 | 0.346 |
| **Interference** | **1.10** | **0.032** | **1.03 – 1.17** | **2.859** | **0.004** |
| **Delay** | **0.83** | **0.032** | **0.78 – 0.88** | **-5.860** | **<0.001** |
| Balance | 1.16 | 0.096 | 0.96 – 1.40 | 1.543 | 0.123 |
| Group * Interference | 1.04 | 0.032 | 0.98 – 1.11 | 1.293 | 0.196 |
| Group * Delay | 0.98 | 0.032 | 0.92 – 1.04 | -0.736 | 0.462 |
| Interference * Delay | 1.00 | 0.032 | 0.94 – 1.07 | 0.033 | 0.974 |
| Group * Balance | 1.13 | 0.096 | 0.93 – 1.36 | 1.253 | 0.210 |
| Interference * Balance | 1.02 | 0.032 | 0.96 – 1.09 | 0.709 | 0.479 |
| Delay * Balance | 1.04 | 0.032 | 0.98 – 1.11 | 1.188 | 0.235 |
| Group * Interference * Delay | 1.05 | 0.032 | 0.98 – 1.11 | 1.415 | 0.157 |
| Group * Interference * Balance | 1.04 | 0.032 | 0.98 – 1.11 | 1.192 | 0.233 |
| Group * Delay * Balance | 0.94 | 0.032 | 0.89 – 1.00 | -1.821 | 0.069 |
| Interference * Delay * Balance | 0.97 | 0.032 | 0.91 – 1.03 | -1.091 | 0.275 |
| Group * Interference * Delay * Balance | 0.95 | 0.032 | 0.89 – 1.01 | -1.643 | 0.100 |
| Observations | 9970 | | | | |
| Marginal R2 / Conditional R2 | 0.027 / 0.180 | | | | |

### Imaging data analysis including control conditions

For the sake of transparency, we conducted the imaging analysis as described in our pre-registration. Results look pretty comparable to what we find in our analysis reported above. Update contrasted with its control condition ("control short) elicits activity in the left putamen, thalamus as well as middle frontal and parietal areas. There was no activity in the right putamen, however. Ignore vs. its temporal control (control long) elicited activity in the occipital lobe, insula, thalamus, inferior parietal, and frontal gyri, as well as in the cingulate and middle frontal gyrus. Contrasting these two contrasts, led to activation mainly in the occipital lobe and in small clusters in the superior and middle frontal gyrus. Similar to our main analysis, this analysis did not reveal any group differences. See Table S4. for all significant clusters and statistical values.

**Table S4.** Significant clusters for the contrasts condition of interest minus control

| contrast | cluster extent | p (FWE-corr) | T | MNI coordinates | | |
|---|---|---|---|---|---|---|
| | | | | x | y | z |
| update > ctrl_short | 502 | 0 | 9.54 | -44 | -46 | 44 |
| | 398 | 0 | 9.4 | 48 | -40 | 48 |
| | 86 | 0 | 6.44 | -46 | 4 | 48 |
| | 23 | 0 | 6.25 | 34 | 20 | 2 |
| | 10 | 0 | 5.95 | -24 | 0 | 66 |
| | 27 | 0.001 | 5.65 | -2 | 22 | 44 |
| | 5 | 0.003 | 5.4 | 28 | 2 | 62 |
| | 4 | 0.004 | 5.32 | -32 | 20 | 2 |
| | 15 | 0.008 | 5.14 | -16 | -2 | 16 |
| | 3 | 0.023 | 4.86 | -26 | -60 | 46 |
| | 3 | 0.028 | 4.79 | 30 | -56 | 42 |
| | 1 | 0.034 | 4.75 | -20 | 2 | 58 |
| | 2 | 0.034 | 4.75 | -24 | 0 | 54 |
| | 1 | 0.036 | 4.73 | 2 | 12 | 58 |
| | 1 | 0.047 | 4.66 | -16 | -6 | 14 |
| ignore > ctrl_long | 13055 | 0 | 22.08 | 12 | -90 | -2 |
| | 82 | 0 | 15.35 | -20 | -30 | -2 |
| | 39 | 0 | 15.15 | 22 | -28 | -2 |
| | 3186 | 0 | 14.45 | -2 | 12 | 50 |
| | 379 | 0 | 12.74 | 32 | 0 | 52 |
| | 164 | 0 | 11.01 | 34 | 20 | 2 |
| | 7 | 0 | 9.89 | -10 | -70 | -14 |
| | 465 | 0 | 9.42 | 44 | 8 | 28 |
| | 571 | 0 | 9.23 | -32 | 20 | 2 |
| | 2 | 0 | 8.53 | 34 | 42 | 24 |
| | 125 | 0 | 7.52 | -46 | 34 | 18 |
| | 31 | 0 | 6.96 | 6 | 4 | 28 |
| | 40 | 0 | 6.7 | 44 | 40 | 14 |
| | 18 | 0 | 6.64 | 8 | -28 | -2 |
| | 21 | 0 | 5.87 | 18 | 8 | 10 |
| | 1 | 0.011 | 5.05 | 16 | -24 | -6 |
| [ignore - ctrl_long] > [update - ctrl_short] | 11262 | 0 | 19.17 | -14 | -84 | -10 |
| | 39 | 0 | 14.48 | 22 | -28 | -2 |
| | 82 | 0 | 14.36 | -20 | -28 | -2 |
| | 2490 | 0 | 8.57 | -4 | 10 | 50 |
| | 6 | 0 | 8.51 | -10 | -70 | -14 |
| | 203 | 0 | 7.22 | 32 | 0 | 52 |
| | 112 | 0 | 6.07 | 42 | 8 | 24 |
| | 37 | 0 | 6 | 54 | 0 | 38 |
| | 21 | 0.004 | 5.31 | 36 | 14 | 4 |
| | 1 | 0.004 | 5.27 | 16 | -24 | -6 |

**Exploratory psychophysiological interaction (PPI) analysis**

Because we speculated that the interaction between condition and beta in the putamen, might be due to top-down PFC interaction, we further decided to run an additional exploratory psychophysiological interaction (PPI) analysis. The aim of this analysis was to look at condition-specific interaction between putamen and PFC.

We employed a generalized PPI approach, as described in Kuhnke, Kiefer & Hartwigsen (2021). We defined the two putamen clusters identified in the above analysis as seed regions. In order to extract subject-specific activation time series of these two group-based seeds, we employed a dynamic approach where individual seed ROIs were defined as the 10% most active voxels for the conceptual contrast within the respective overall group seed mask. After that, individual subject data were modeled separately using the gPPI toolbox (version 13.1; https://www.nitrc.org/projects/gppi). This first-level GLM included: 1) "Psychological" regressors for each task event, that is, stick functions at event onsets convolved with the canonical hemodynamic response function; 2) A "physiological" regressor formed by the first eigenvariate of the respective seed ROI time series; and 3) PPI regressors for each task event created by multiplying the deconvolved BOLD signal of the seed ROI with the condition onsets and convolving with the canonical HRF (Gitelman et al. 2003; McLaren et al. 2012). Finally, we also included all six nuisance regressors i.e. the six head motion parameters. Contrast images were computed for each participant and submitted to t-tests at the group level. To test for functional coupling at the time of ignore vs. update, we compared the interaction of putamen and other areas at the time of ignore minus update. To investigate if any potential interactions would be behaviorally relevant, we also added condition-specific accuracy as a covariate of interest. We did not include the factor group, since our previous analyses point towards no group differences. All activation maps were thresholded at a cluster-wise $p < 0.05$ family-wise error (FWE) corrected for multiple comparisons.

Results showed that for the right as well as left putamen seed there were significant interactions with the thalamus, the (rest of the) putamen, the parietal lobe, as well as frontal areas. See Figure S2 for a visual overview and Table S5 for all cluster statistics. There was no difference among conditions for these interactions, however (i.e. there were no significant PPI clusters when looking at the contrast ignore minus update). Moreover, there was no significant interaction with behavior.

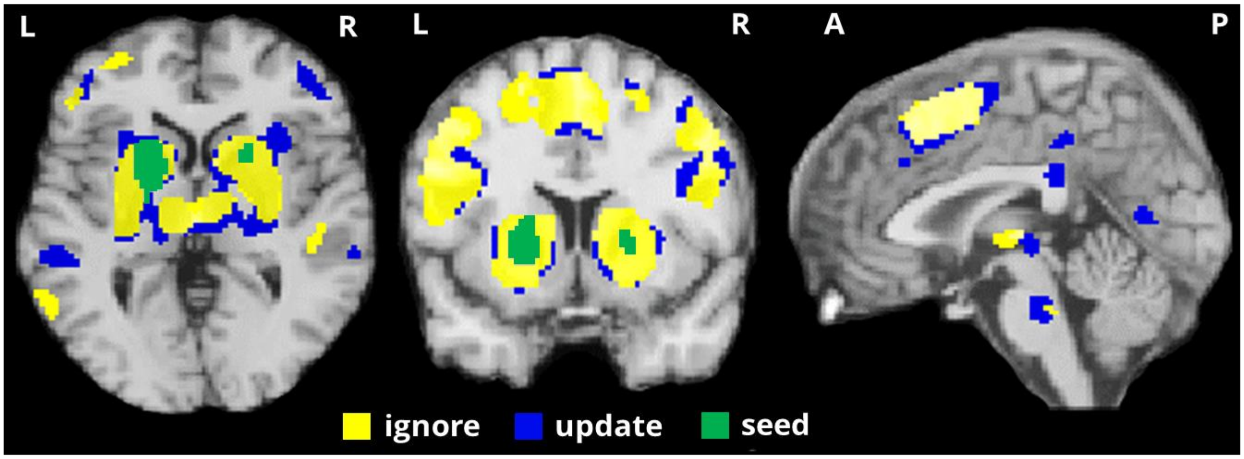


**Figure S2.** Significant clusters resulting from PPI analysis. Activation of (combined) left and right putamen seeds (green) interact with thalamus, parietal lobe, and frontal areas during ignore (yellow) and update (blue).

**Table S4** Significant clusters for psychophysiological interaction with putamen seed

| seed | condition | cluster extent | *p* (FWE-corr) | T | MNI coordinates x | y | z |
|---|---|---|---|---|---|---|---|
| putamen right | ignore | 5478 | 0 | 16.19 | 22 | 10 | 6 |
| | | 223 | 0 | 7.02 | -46 | 34 | 16 |
| | | 233 | 0 | 6.43 | 32 | -42 | 42 |
| | | 310 | 0 | 6.37 | -50 | 8 | 24 |
| | | 92 | 0 | 6.07 | 48 | 28 | -10 |
| | | 472 | 0 | 6.02 | -4 | 0 | 62 |
| | | 104 | 0 | 5.85 | -48 | -6 | 46 |
| | | 76 | 0 | 5.82 | 26 | -8 | 50 |
| | | 113 | 0 | 5.79 | 56 | -10 | -4 |
| | | 74 | 0 | 5.6 | -58 | -38 | 8 |
| | | 26 | 0.005 | 5.54 | 58 | -18 | 30 |
| | | 88 | 0 | 5.53 | 44 | 2 | 44 |
| | | 134 | 0 | 5.46 | 40 | 28 | 16 |
| | | 10 | 0.015 | 5.42 | -4 | -28 | -26 |
| | | 109 | 0 | 5.38 | -46 | -36 | 48 |
| | | 12 | 0.013 | 5.35 | -50 | 36 | 4 |
| | | 9 | 0.016 | 5.31 | 30 | 36 | -12 |
| | | 1 | 0.039 | 5.3 | -38 | 2 | -40 |
| | | 39 | 0.002 | 5.3 | -62 | -56 | 2 |
| | | 13 | 0.012 | 5.29 | -30 | -46 | 50 |
| | | 19 | 0.008 | 5.22 | -20 | -2 | 54 |
| | | 31 | 0.004 | 5.18 | -48 | -18 | -8 |
| | | 2 | 0.033 | 5.15 | -16 | -58 | -34 |
| | | 12 | 0.013 | 5.13 | -52 | -48 | 40 |
| | | 8 | 0.018 | 5.11 | -4 | 30 | 50 |
| | | 18 | 0.008 | 5.07 | 48 | -28 | 0 |
| | | 3 | 0.029 | 5.02 | 42 | 18 | 18 |
| | | 3 | 0.029 | 4.94 | 54 | -22 | 40 |
| | | 3 | 0.029 | 4.93 | -2 | 38 | 28 |
| | | 1 | 0.039 | 4.92 | -18 | 12 | 60 |
| | update | 1803 | 0 | 14.2 | 20 | 8 | 6 |
| | | 3086 | 0 | 11.56 | -18 | 10 | 4 |
| | | 655 | 0 | 7.74 | -46 | -6 | 46 |
| | | 3245 | 0 | 7.1 | -6 | 4 | 60 |
| | | 257 | 0 | 6.6 | 60 | -26 | -4 |
| | | 744 | 0 | 6.5 | -52 | 24 | 14 |
| | | 459 | 0 | 6.04 | -50 | -48 | 44 |
| | | 55 | 0.001 | 5.96 | -6 | -28 | 40 |
| | | 197 | 0 | 5.81 | -54 | -42 | 12 |
| | | 27 | 0.005 | 5.56 | -10 | -30 | -34 |
| | | 109 | 0 | 5.53 | -50 | -36 | 0 |
| | | 19 | 0.007 | 5.53 | 48 | -42 | 8 |
| | | 22 | 0.006 | 5.49 | -2 | -24 | -26 |
| | | 19 | 0.007 | 5.4 | 42 | 52 | 4 |
| | | 21 | 0.007 | 5.33 | -40 | -48 | -18 |
| | | 29 | 0.004 | 5.3 | 6 | 24 | 32 |
| | | 15 | 0.01 | 5.27 | -2 | 42 | 26 |
| | | 5 | 0.023 | 5.11 | 8 | -14 | 42 |
| | | 6 | 0.021 | 5.1 | -2 | -66 | 10 |
| | | 9 | 0.016 | 5.08 | -18 | 10 | 58 |
| | | 4 | 0.026 | 5.07 | 48 | -44 | 22 |
| | | 11 | 0.013 | 5.06 | 48 | 34 | 16 |
| | | 5 | 0.023 | 5.05 | 34 | 50 | 22 |
| | | 8 | 0.017 | 5 | 48 | 38 | -4 |
| | | 1 | 0.038 | 4.95 | 6 | -36 | 46 |

**Table S4** Significant clusters for psychophysiological interaction with putamen seed

| seed | condition | cluster extent | *p* (FWE-corr) | T | MNI coordinates | | |
|---|---|---|---|---|---|---|---|
| | | | | | x | y | z |
| putamen left | ignore | 5238 | 0 | 12.34 | -24 | -4 | 6 |
| | | 4897 | 0 | 8 | -46 | 34 | 16 |
| | | 1806 | 0 | 7.49 | -46 | -40 | 46 |
| | | 1930 | 0 | 7.21 | 36 | 0 | 48 |
| | | 1180 | 0 | 7.16 | 40 | -44 | 50 |
| | | 141 | 0 | 6.18 | -60 | -58 | 6 |
| | | 67 | 0.001 | 5.83 | -2 | -32 | -30 |
| | | 27 | 0.006 | 5.4 | -64 | -40 | -10 |
| | | 35 | 0.004 | 5.33 | 56 | -6 | -12 |
| | | 21 | 0.008 | 5.22 | -38 | -52 | -12 |
| | | 10 | 0.017 | 5.21 | -44 | -50 | -22 |
| | | 29 | 0.005 | 5.2 | 48 | -28 | 2 |
| | | 3 | 0.031 | 5.1 | 48 | 44 | -6 |
| | | 8 | 0.02 | 5.1 | -8 | -46 | 44 |
| | | 8 | 0.02 | 5.1 | 8 | -22 | -28 |
| | | 2 | 0.035 | 5.06 | 46 | 30 | 38 |
| | | 6 | 0.023 | 5.01 | 14 | -26 | 64 |
| | | 2 | 0.035 | 4.93 | -10 | -30 | 60 |
| | | 2 | 0.035 | 4.91 | 20 | -24 | -2 |
| | | 2 | 0.035 | 4.9 | -66 | -34 | 0 |
| | | 2 | 0.035 | 4.88 | 14 | -64 | 54 |
| | update | 11731 | 0 | 13.13 | -18 | 10 | 4 |
| | | 5224 | 0 | 8.19 | 52 | -26 | 46 |
| | | 2177 | 0 | 7.19 | -50 | -30 | 44 |
| | | 276 | 0 | 6.29 | -6 | -30 | -30 |
| | | 26 | 0.006 | 5.99 | -16 | -62 | -32 |
| | | 81 | 0.001 | 5.87 | -48 | -44 | -18 |
| | | 40 | 0.003 | 5.79 | -34 | 34 | 44 |
| | | 207 | 0 | 5.79 | -56 | -34 | 0 |
| | | 292 | 0 | 5.75 | -10 | -76 | 10 |
| | | 200 | 0 | 5.71 | -58 | -48 | 18 |
| | | 29 | 0.005 | 5.56 | 30 | -58 | -8 |
| | | 12 | 0.015 | 5.49 | 66 | -38 | -8 |
| | | 152 | 0 | 5.48 | 54 | -30 | -2 |
| | | 12 | 0.015 | 5.46 | -28 | -8 | -32 |
| | | 35 | 0.004 | 5.31 | -46 | -78 | 12 |
| | | 21 | 0.008 | 5.29 | 48 | -74 | 14 |
| | | 8 | 0.02 | 5.29 | 38 | -32 | -22 |
| | | 25 | 0.007 | 5.21 | -22 | 20 | 46 |
| | | 31 | 0.005 | 5.19 | 58 | -44 | 26 |
| | | 12 | 0.015 | 5.17 | -28 | -60 | -12 |
| | | 3 | 0.031 | 5.13 | 26 | -20 | -26 |
| | | 19 | 0.009 | 5.09 | 14 | -66 | 20 |
| | | 4 | 0.028 | 5.09 | 48 | -18 | -8 |
| | | 9 | 0.018 | 5.06 | -8 | -48 | 46 |
| | | 6 | 0.023 | 5.06 | -12 | -32 | 68 |
| | | 7 | 0.021 | 5.05 | 62 | -10 | -14 |
| | | 7 | 0.021 | 5.03 | 8 | -52 | 54 |
| | | 1 | 0.04 | 5.02 | -30 | 4 | -36 |
| | | 2 | 0.035 | 4.99 | -40 | 0 | -38 |
| | | 4 | 0.028 | 4.97 | 32 | 10 | -36 |
| | | 1 | 0.04 | 4.9 | 46 | -40 | 8 |
| | | 1 | 0.04 | 4.87 | -28 | 0 | -36 |
